# Evaluating the implementation of weekly rifapentine-isoniazid (3HP) for tuberculosis prevention among people living with HIV in Uganda: A qualitative evaluation of the 3HP Options Trial

**DOI:** 10.1101/2024.08.19.24308041

**Authors:** Allan Musinguzi, Joan R. Kasidi, Jillian L. Kadota, Fred Welishe, Anne Nakitende, Lydia Akello, Jane Nakimuli, Lynn T. Kunihira, Bishop Opira, Yeonsoo Baik, Devika Patel, Amanda Sammann, Christopher A. Berger, Hélène E. Aschmann, Payam Nahid, Robert Belknap, Moses R. Kamya, Margaret A. Handley, Patrick PJ Phillips, Noah Kiwanuka, Achilles Katamba, David W. Dowdy, Adithya Cattamanchi, Fred C. Semitala, Anne R. Katahoire

**Author notes:** Corresponding author (FCS). Contributed equally as first authors (Allan Musinguzi, Joan R. Kasidi). Contributed equally as senior authors (Fred C. Semitala, Anne R. Katahoire). **Trial registration:** ClinicalTrials.gov (NCT03934931); Registered 2^nd^ May 2019; https://clinicaltrials.gov/study/NCT03934931?id=NCT03934931&rank=1.

## Abstract

Three months of isoniazid-rifapentine (3HP) is being scaled up for tuberculosis (TB) preventive treatment (TPT) among people living with HIV (PLHIV) in high-burden settings. More evidence is needed to identify factors influencing successful 3HP delivery. We conducted a qualitative assessment of 3HP delivery nested within the 3HP Options Trial, which compared three optimized strategies for delivering 3HP: facilitated directly observed therapy (DOT), facilitated self-administered therapy (SAT), and patient choice between facilitated DOT and facilitated SAT at the Mulago HIV/AIDS clinic in Kampala, Uganda. We conducted 72 in-depth interviews among PLHIV purposively selected to investigate factors influencing 3HP acceptance and completion. We conducted ten key informant interviews with healthcare providers (HCPs) involved in 3HP delivery to identify facilitators and barriers at the clinic level. We used post-trial 3HP delivery data to assess sustainability. We conducted an inductive thematic analysis and aligned the emergent themes with the RE-AIM framework dimensions to report implementation outcomes. Understanding the need for TPT, once-weekly dosing, shorter duration, and perceived 3HP safety enhanced acceptance overall. Treatment monitoring by HCPs and reduced risk of HIV status disclosure enabled DOT acceptance. Dosing autonomy enabled SAT acceptance. Switching between DOT and SAT as required enabled acceptance for patient choice. Dosing reminders, reimbursement for clinical visits, and social support enabled 3HP completion; pill burden, side effects, and COVID-19-related treatment restrictions hindered completion. All HCPs were trained and participated in 3HP delivery with high fidelity. Training, care integration, and collaboration among HCPs enabled, whereas initial concerns about 3HP safety among HCPs delayed 3HP adoption and implementation. SAT was maintained post-trial; DOT was discontinued due to inadequate ongoing financial support beyond the study period. Facilitated delivery strategies made 3HP treatment convenient for PLHIV and were feasible and implemented with high fidelity by HCPs. However, the costs of 3HP facilitation may limit wider scale-up.

## Introduction

Scaling up short-course tuberculosis (TB) preventive treatment (TPT) regimens is key to achieving ambitious global targets to end TB, especially among people living with HIV (PLHIV) in high-burden settings (1, 2). Newer TPT regimens, including weekly isoniazid and rifapentine for three months (3HP), have well-documented advantages (higher tolerability and completion) over the traditional six to nine months regimen of daily isoniazid (isoniazid preventive therapy, IPT) (3, 4) and are recommended by the World Health Organization (WHO) (5). However, the best approach to delivering 3HP to PLHIV in high-burden TB/HIV settings remains unclear. Although effective (3, 6), 3HP delivery by directly observed therapy (DOT) may not be cost-effective in high-burden, low-income settings (7, 8). Self-administered therapy (SAT) overcomes most of the barriers associated with DOT but may be less effective in high-burden settings (9).

Therefore, to identify the optimal approach to 3HP delivery for PLHIV in a high-burden TB/HIV setting, we conducted a pragmatic randomized implementation trial (3HP Options Trial) of 3HP delivery strategies (facilitated DOT, facilitated SAT, or providing an informed choice between facilitated SAT and DOT using a shared decision aid) among 1655 PLHIV (with equal participant allocation per delivery strategy) at a high volume, urban HIV clinic in Kampala, Uganda, from July 13, 2020 to July 8, 2022 (10). The facilitated 3HP delivery strategies were optimized to promote facilitators (fear of contracting TB, trust in healthcare providers, and perceived benefits of DOT and SAT) and overcome the important barriers (lack of knowledge about TB/TPT, pill burden, potential side effects of TPT, and the perceived difficulties of DOT and SAT) identified through formative qualitative research (11). The 3HP Options Trial demonstrated >90% acceptance and completion of 3HP for all three delivery strategies, with no significant differences between strategies. Overall, <1% of trial participants experienced an adverse event requiring treatment discontinuation (12, 13).

Here, guided by the RE-AIM implementation science framework (14, 15), we conducted an explanatory qualitative evaluation (16) to understand the processes and contextual factors that influenced 3HP acceptance and completion overall and within each delivery strategy during the trial; the clinic-level facilitators and barriers to adoption of the facilitated 3HP delivery strategies; the implementation of 3HP under each delivery strategy; and the sustainability of 3HP delivery at the trial site. By focusing on the perspectives of PLHIV who were offered 3HP, healthcare providers (HCPs) who provided the services, and the clinical and socioeconomic context in which the services were provided, we aimed to examine factors likely to enable or hinder the integration of 3HP into policy and practice.

## Materials and Methods

### Study design

This was an explanatory qualitative study in which we evaluated the implementation of 3HP during the 3HP Options Trial (10) in Kampala, Uganda. We assessed the reach, effectiveness, adoption, implementation, and maintenance domains of the RE-AIM framework. Detailed descriptions of the RE-AIM framework, its dimensions, and its application to this study are shown in **Table 1**. We followed the Consolidated Criteria for Reporting Qualitative Research (COREQ) when writing this manuscript (17). This study was approved by the School of Public Health Research Ethics Committee at the Makerere University College of Health Sciences (Kampala, Uganda), the Uganda National Council for Science and Technology (Kampala, Uganda), and the University of California San Francisco Institutional Review Board (San Francisco, CA, USA).

**Table 1.**
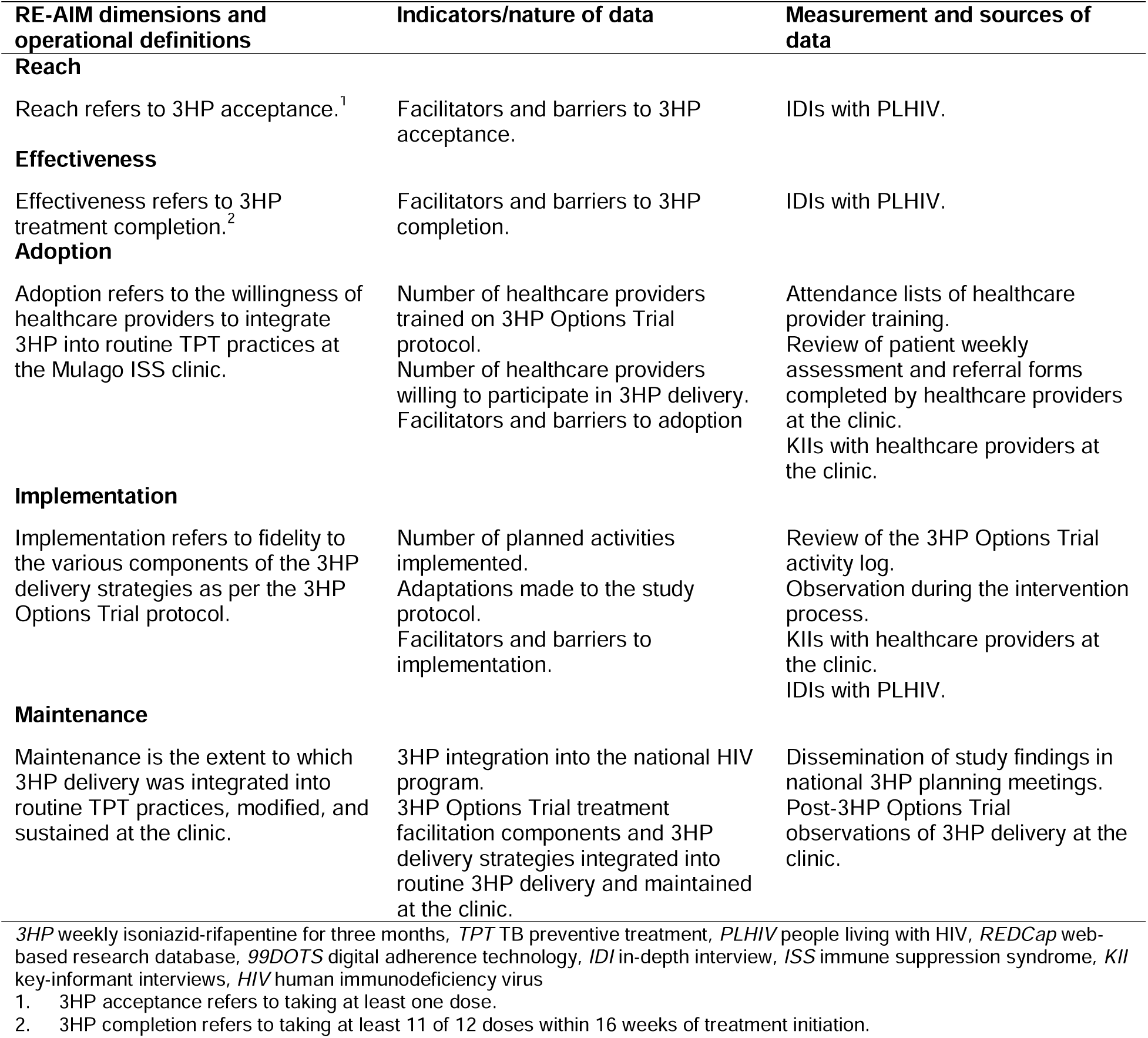
The RE-AIM framework adapted to the evaluation of 3HP delivery in the 3HP Options Trial.

### Study setting

The research was conducted at the Mulago Immune Suppression Syndrome (ISS) clinic run by the Makerere University Joint AIDS Program (MJAP) in Kampala, Uganda. This clinic is the largest specialized outpatient HIV clinic in Uganda, providing comprehensive HIV/AIDS care and treatment services to more than 16,000 clients at no cost. The clinic started offering IPT to eligible PLHIV in 2017. However, 3HP was unavailable at the clinic through the national HIV program until July 2022.

### Implementation strategies

The 3HP Options trial evaluated three pragmatic 3HP delivery strategies: facilitated DOT, facilitated SAT, and informed patient choice between facilitated DOT and facilitated SAT (with the help of a decision aid). Details of these strategies are described elsewhere (10). Briefly, each strategy included standardized pre-treatment counseling, streamlined clinic visits, dosing reminders via the 99DOTS digital adherence technology platform from Bengaluru, India (18), and transport reimbursement (∼4-8 USD/visit) per clinic visit. DOT participants took all 12 weekly 3HP doses under the direct observation of HCPs at the clinic. SAT participants were required to take doses one, six, and 12 under direct observation at the clinic. They were provided with a waterproof pill pack containing pre-packaged doses 2-5 and 7-11, along with a card insert that had a toll-free telephone number to confirm their weekly dosing from home. Participants in the informed patient choice strategy were given the option to switch between DOT and SAT as required during 3HP treatment. Prior to the recruitment of study participants, all HCPs at the clinic received training on delivering and managing 3HP treatment and addressing any adverse events. Side effects and adherence monitoring were done by HCPs during weekly clinic visits for DOT and via the 99DOTS platform for SAT.

### Study population

We interviewed a subset of PLHIV and HCPs for the qualitative research. PLHIV were approached either in person during clinic visits or via telephone calls during the COVID-19 pandemic. All PLHIV provided written informed consent to future selection for participation in in-depth interviews (IDIs) at enrollment in the main trial, and additional verbal consent was sought for IDIs conducted over the telephone. We selected equal numbers of PLHIV per 3HP delivery strategy for inclusion in the IDIs based on age, sex, duration of antiretroviral therapy (ART), 3HP treatment outcome, and 99DOTS engagement. Given that we were studying a single-center homogeneous population, a maximum sample size of 17 IDIs per 3HP delivery strategy would have sufficed to achieve thematic saturation, according to studies of empirical data (19). Every three months, during the two years of participant recruitment for the main trial, we selected three PLHIV enrolled in that period, per delivery strategy, for a pre-determined sample size of 72 IDI participants (24/delivery strategy).

HCPs were selected for inclusion in key-informant interviews (KIIs) based on their cadres and active involvement in implementing the 3HP Options Trial at the clinic. HCPs were approached in person and provided written informed consent to participate. The number of HCPs directly involved in 3HP implementation determined the KII sample size.

### Data collection and management

We conducted IDIs and KIIs between 27^th^ November 2020 and 30^th^ March 2022. We developed interview guides based on our research questions and pretested them with PLHIV and HCPs at the clinic. JRK, a trained female social science researcher who was not familiar with study participants, conducted all the interviews. Prior to each interview, JRK established rapport with the participant and shared interview objectives. Interviews were conducted in person at the clinic in a private area with only the participant and interviewer in the room. Due to local travel restrictions during the COVID-19 pandemic, interviews were conducted over the telephone while respecting participants’ privacy. Each interview lasted 35-50 minutes. IDIs with PLHIV explored contextual and process factors that influenced 3HP acceptance and completion. We defined 3HP acceptance as taking at least one dose and completion as taking at least 11 of 12 doses within 16 weeks of treatment initiation. We defined contextual factors as those related to the settings where the PLHIV lived, worked, and within which 3HP implementation occurred. We defined process factors as those related to the 3HP TPT regimen and its delivery per the study protocol.

HCP interviews explored their perspectives on implementing 3HP in the clinic using the three delivery strategies. The interviews further explored changes made in the patient workflow in the clinic during the implementation of the 3HP delivery strategies. KIIs were conducted towards the end of participant recruitment in the main trial to facilitate a better evaluation of the study. We achieved data saturation based on meaning saturation for IDIs and KIIs when no new details/aspects were identified for the various emergent codes (20).

All interviews were audio-recorded and transcribed verbatim. Expert translation was used to convert Luganda transcripts to English. Prior to importing transcripts into NVivo V1.6.1 (21), we ensured their accuracy and anonymity.

Finally, we conducted post-trial observations of 3HP delivery at the clinic for 14 months (November 2022 to December 2023) to assess the continuity of the study interventions.

### Data analysis

The analytical process was led by a doctoral-trained social and behavioral scientist employed as a Professor at Makerere University, Kampala (ARK, female). It involved ARK and JRK reading and re-reading transcripts and open-coding the data. Weekly reflection meetings with ARK, JRK, AM, and FCS involved discussions, questions, and reflections on the coding process, resulting in valuable feedback and consensus. This collaborative process achieved the final coding and theme development, which enhanced reflexivity and interpretative depth. The codes and themes generated through the inductive process were mapped onto the five dimensions of the RE-AIM framework (reach, effectiveness, adoption, implementation, and maintenance) (14, 15, 22). Corresponding quotations were extracted from the transcripts. The “reach” dimension was used to reflect 3HP acceptance, whereas the “effectiveness” dimension reflected completion. The “adoption,” “Implementation,” and “maintenance” dimensions were used to reflect adoption, implementation, and sustainability, respectively, of 3HP delivery strategy components. Both inductive and deductive thematic analyses were used to interpret the data, aligning the emergent themes to a pre-existing theoretical framework.

### Data validation and feedback to study participants and stakeholders

We held two validation meetings with HCPs at the clinic, shared our findings, and received feedback. We also shared the study results with PLHIV at the clinic during routine health education talks. HCPs and PLHIV confirmed that the findings resonated with their experiences. We shared our findings with the Ministry of Health in Uganda through the National TB and Leprosy Program’s 3HP planning meetings before the HIV program roll-out of 3HP in Uganda.

## Results

### Characteristics of interview participants

Seventy-two PLHIV (24 per 3HP delivery strategy) participated in the IDIs. Of these, 42 (58%) were female, and the median age was 40 years (interquartile range [IQR]: 30.5-45). The median time on ART was 6.9 years (IQR: 1.9-11.3), and 17 (24%) reported prior TB disease. Ten healthcare providers participated in the KIIs. Of these, eight (80%) were male, and the median age was 31.5 years (IQR: 29-36). The duration in service at the current post ranged from six months to 17 years (median 4 years; IQR: 3-7).

### Reach

#### Facilitators of reach

The high overall acceptance of 3HP treatment was attributed to three main factors. First, following pre-treatment counseling, PLHIV perceived themselves as being at a high risk of contracting TB. Second, the shorter duration and once-a-week 3HP dosing schedule were convenient for them. Finally, PLHIV perceived 3HP to be safe and tolerable, which was reinforced by positive feedback from both HIV peers and HCPs at the clinic. **Table 2** summarizes the facilitators of reach reported by PLHIV, overall and within each 3HP delivery strategy:

> “I didn’t fear, because at first, I had feared, but when I saw others going for it, others were saying it doesn’t treat them badly, I said now for me why do I fear?” (Male participant, Facilitated SAT).

**Table 2.**
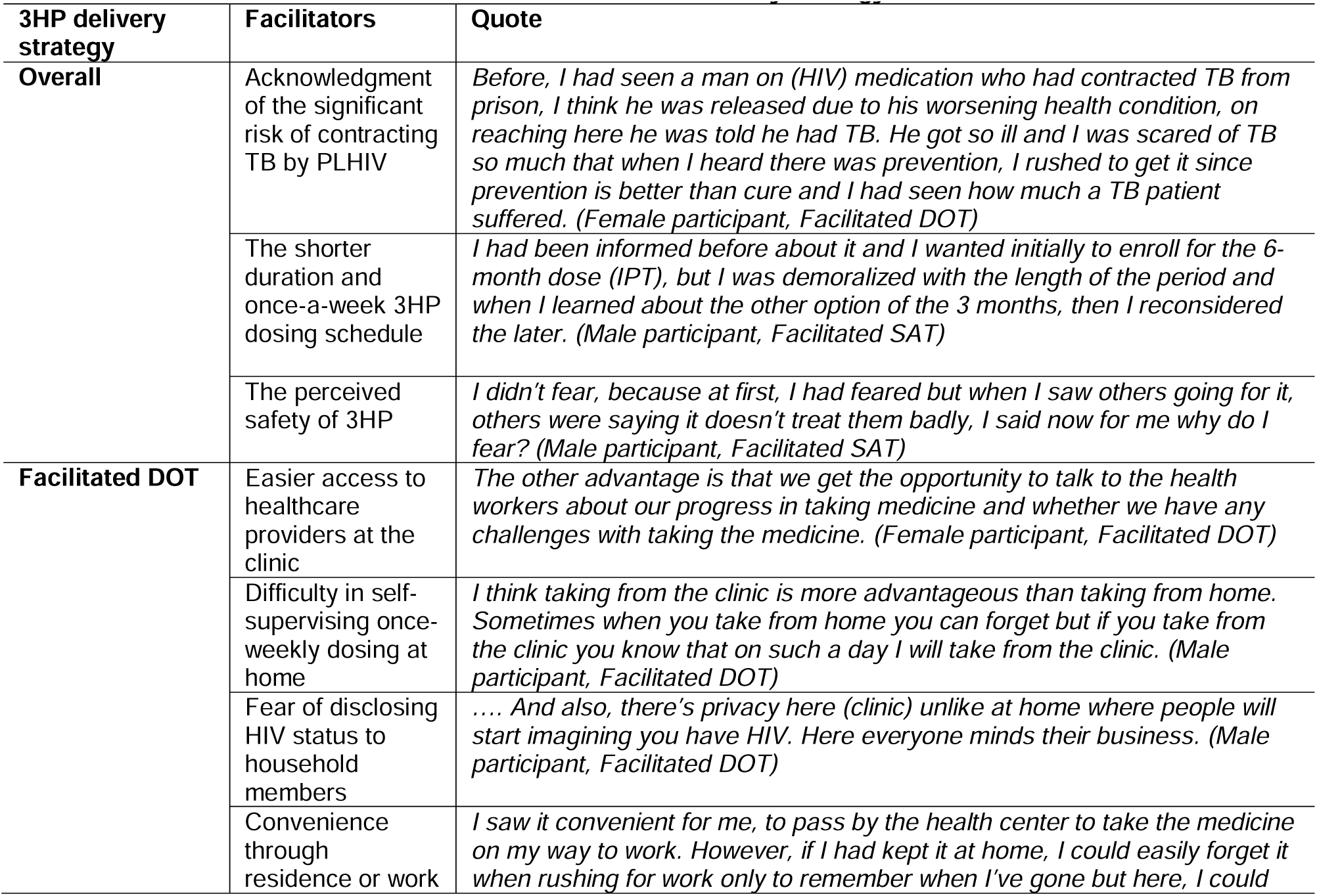
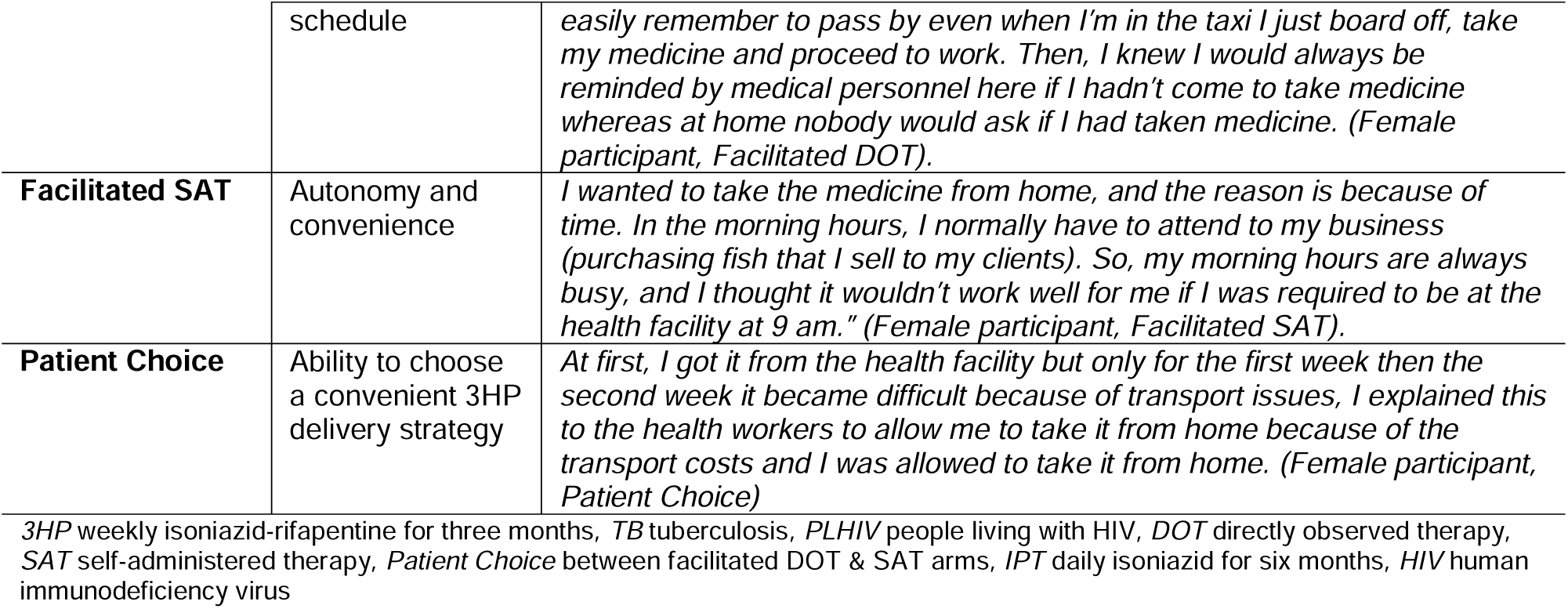
Facilitators of reach overall and within each 3HP delivery strategy.

Most participants who received 3HP under DOT accepted treatment because DOT was better aligned with their lifestyle and addressed their perceived barriers to treatment, including the fear of accidentally disclosing their HIV status to significant others or other household members if they had to take their medicines from home. This was especially true among young adults:

> “You see at home it would have been very difficult for me because I share (a room) with a friend. The reason is that he would not have liked it maybe or he would have viewed it negatively. The other thing is that it becomes difficult to hide.” (Male participant, Patient Choice).

Furthermore, PLHIV for whom the clinic was nearby either through residence or work and those who preferred the real-time, in-person reassurance of a health worker while taking their medicines, either for fear of drug-related side effects or the inability to self-administer the once-weekly 3HP dosing schedule, were more comfortable with DOT:

> “I saw it convenient to pass by the health center to take the medicine on my way to work. However, if I had kept it at home, I could easily forget it when rushing for work only to remember when I’ve gone but here, I could easily remember to pass by even when I’m in the taxi I board off, take my medicine and proceed to work. Then, I knew I would always be reminded by medical personnel here if I hadn’t come to take medicine, whereas at home, nobody would ask if I had taken medicine.” (Female participant, Facilitated DOT).

The convenience and autonomy associated with SAT were the strongest facilitators of treatment acceptance among PLHIV with busy work and daily life schedules:

> “That’s what I wanted because you may have gone for a trip and you find that it will coincide with that Wednesday or you are going on Tuesday it means you have to come on Tuesday now the days don’t connect but if you swallow from home when they give it to you even when you go on Wednesday you go with your medicine.” (Male participant, Facilitated SAT).

One of the main reasons why patients in the informed patient choice group accepted treatment was because they were given the freedom to choose a delivery method that was convenient for them:

> “At first, I got it from the health facility but only for the first week, then the second week, it became difficult because of transport issues; I explained this to the health workers to allow me to take it from home because of the transport costs and I was allowed to take it from home.” (Female participant, Patient Choice).

### Effectiveness

#### Facilitators of effectiveness

PLHIV reported that automated weekly dosing (SAT) or clinic appointment (DOT) reminders coordinated through the 99DOTS digital adherence technology platform helped ensure treatment completion across all delivery strategies. **Table 3** provides a summary of what enabled and hindered treatment effectiveness as reported by participants, both overall and within each 3HP delivery strategy:

> “I used to get a call on Monday reminding me to take my drugs. It was good and very helpful because it reminded me to take my drugs. Anyone can forget.” (Male participant, Facilitated SAT).

**Table 3.** Facilitators and barriers to effectiveness overall and within each 3HP delivery strategy.

| 3HP delivery strategy | Facilitators | Barriers |
| --- | --- | --- |
| Overall | **Automated weekly dosing/clinic appointment reminders** - *It helped me because doctor, you may forget and may not know that tomorrow, I have to return but when they remind you by calling you, you get to know and say eh! Tomorrow I am returning to the hospital. Then you prepare and come. (Female participant, Facilitated DOT)* | **High pill burden at the beginning using loose pills of isoniazid and rifapentine (eleven pills per weekly dose)** - *Oh, the number yes, it was a challenge, they were so many that after taking them you could feel like vomiting. (Female participant, Patient Choice)* |
|  | **Reimbursement of travel costs** - *Another benefit was that we used to come with the hope that I would take my drugs from the clinic but there was something else that I would carry home. That also encouraged us. Personally, it encouraged me so much. (Female participant, Facilitated DOT)* | **3HP-related side effects** - *After the first dose I felt sick until I decided to stop taking the medicine. (Female participant, Facilitated SAT)* |
|  | **Social support** - *The boss (work employer) first asked me to explain to him..... he said if that is it, okay it's good, every Wednesday when it reaches tell me so that if you have a safari to go I can cancel and another one goes or you look for another driver you give him and he goes then you wait for your day. (Male participant, Facilitated SAT)* | **COVID-19 pandemic-related treatment disruptions** - *I stopped during the COVID-19 lockdown because the situation was tough. I had to walk to pick up the medication (from the clinic) given that there were no transportation means.... And because I was not earning, I decided to go to the village. (Female participant, Patient Choice)* |
| Facilitated SAT | **Packaging of 3HP medicines** - *"It's a good one (the pill pack) that it keeps the medication safe. You know, at home, you might have people touching here and there, so once they know that that thing is for your medication, nobody will tamper with it. So, it keeps the medication safe." (Male participant, Facilitated SAT).* |  |

3HP weekly isoniazid-rifapentine for three months, DOT directly observed therapy, SAT self-administered therapy, Patient Choice between facilitated DOT & SAT arms

Reimbursement of participants’ travel costs associated with clinic visits also enabled adherence and completion:

> “The most encouraging thing is that as we left, they would give us a transport refund. So, even if I did not have money on the day of picking medication, I would borrow from someone because I knew I would get a refund…… this helped me a lot.” (Female participant, Facilitated SAT).

Across all delivery strategies, support from family, friends, and, in some cases, workmates motivated participants to complete treatment:

> “I told my mother and my sister. They told me it was a nice initiative, and they used to encourage me to take the medicines because they knew that I had HIV. My mother was ready to help me with transport if I ran short of it.” (Female participant, Facilitated DOT).

Furthermore, among PLHIV who received 3HP under SAT, the labeling and packaging of their medicines enabled correct and timely dosing, as well as safe storage at home:

> “It’s a good one (the pill pack) that it keeps the medication safe. You know, at home, you might have people touching here and there, so once they know that that thing is for your medication, nobody will tamper with it. So, it keeps the medication safe.” (Male participant, Facilitated SAT).

#### Barriers to effectiveness

Some PLHIV had difficulty completing 3HP treatment at the start of the study, before the switch to fixed-dose combination (FDC) pills, due to the high number of loose pills (eleven) in the weekly dose, in all delivery strategies:

> “Oh, the number (of pills), yes, it was a challenge; they were so many that after taking them you could feel like vomiting.” (Female participant, Patient Choice).

In all delivery strategies, treatment-related side effects caused significant discomfort in some cases, affected participant livelihoods, and led to non-adherence and, ultimately, discontinuation of treatment:

> “It was all okay, but the side effects of the drug could not allow me to continue medication because I had decided to take my medicine, I realized I work for myself no one helps me so if I fail to work and all I do is to sleep, who will feed me? And that’s why I stopped taking it.” (Female participant, Facilitated SAT).

The COVID-19 pandemic disrupted participants’ treatment plans across all delivery strategies. Community lockdowns resulted in restricted movement and increased transportation costs for clinic visits. Some PLHIV lost their livelihoods and had to relocate further away from the clinic, making it harder to attend clinic visits. Others feared contracting COVID-19 from the clinic and, therefore did not return for the scheduled visits:

> “Before the lockdown, I had taken the medicine for 6 weeks, and when the lockdown was announced, movement was impossible.” (Female participant, Facilitated DOT).

### Adoption

#### Facilitators of adoption

Since 3HP was a new treatment, a training protocol was established to facilitate clinic staff education. The training was incorporated into the clinic’s routine weekly continuing medical education (CME) sessions. A total of 87 clinic staff members were trained, including 14 HCPs who were directly involved in the delivery of 3HP according to the study protocol.

> “…. The most important thing is the education. If there is continuous education, then things become easy. So, we had quite a number of CMEs before the real implementation of 3HP and people got to know what 3HP is; what it involves; who can get 3HP and who cannot.” (Male HCP, interview 06)

In addition to the general 3HP protocol training, the staff who directly participated in 3HP delivery were provided with job aids and standard operating procedures (SOPs) highlighting the management of common 3HP-related side effects.

> “We had training, and then they gave us job aids. We were given some books (3HP treatment protocols and standard operating procedures). These told us who qualifies for 3HP, how to grade adverse drug reactions, how to manage the different adverse drug reactions, and how to fill out the ADR (adverse drug reaction) form. So, they took us through all those phases. Then still, study staff used to come during the introductory phases to guide us on what to do and what not to do because initially, we used to get scared and stop drugs even on a minor adverse drug reaction. So, they would come and guide us.” (Female HCP, interview 03).

Three refresher training sessions were conducted during the 3HP treatment phase of the study to address ongoing implementation challenges and provide healthcare providers with updates about the project.

> “Midway, there was refresher training about the study, so I feel that should stay…. There were updates about the project. Some patients had developed side effects, but most of them were doing well with 3HP on a large scale. New information should always be passed on to the doctors to ensure they don’t forget the information they learned earlier.” (Male HCP, interview 01).

### Implementation

#### Fidelity of implementing 3HP delivery

3HP delivery was implemented as planned. However, there were two important protocol adaptations, including a switch to the 3HP fixed-dose combination and an increase in participants’ transport reimbursement.

#### Switch to the 3HP fixed-dose combination

When participant recruitment began on July 13th, 2020, the only form of the 3HP regimen available was loose pills containing rifapentine (150mg) and isoniazid (300mg). The weekly dose of 900mg required taking eleven pills, including two 25mg pyridoxine pills. However, in September 2020, Macleods Pharmaceuticals Limited released the 3HP FDC pills, which contain rifapentine and isoniazid in 300mg/300mg doses. On November 2nd, 2020, we began offering the 3HP-FDC pills after receiving the necessary regulatory approvals. With this new option, PLHIV no longer needed to take eleven pills per week, but only five. Some expressed difficulty in swallowing the large FDC pills, but most PLHIV appreciated the decrease in the number of pills.

> “It changed, the last dose changed…. they gave me one tablet that had a combination of very many. I took one tablet, it was big. I can’t remember what the combination was but I know there was one tablet which they gave me which was big…. I prefer this one; the one tablet…eh, eh, eh, the other ones (initial eleven pills) were too many.” (Female participant, Facilitated SAT)

#### Increase in participant travel cost reimbursement

Due to COVID-19-related travel restrictions and the resulting rise in public transportation costs, we increased participant travel cost reimbursement from 4 to 8 USD to facilitate treatment completion. This also enabled PLHIV, who had to miss work, to attend clinic visits to adhere to and complete treatment.

> “I was not affected because they gave us transport. So, you would know that even if you missed work, the money would help with transport and meals for the day depending on the circumstances…. The first day, they gave me 15,000 (Uganda) shillings; the second time, I got 15,000. The following days, when I came, they gave me 30,000 shillings. It was enough. (Female participant, Facilitated DOT)

#### Facilitators of implementation

The clinic had already been involved in preventing, diagnosing, and treating TB even before the introduction of the 3HP treatment. This reportedly made the implementation process of 3HP easier. PLHIV were already receiving daily IPT for six months, and there were already existing SOPs in place for TB prevention, diagnosis, and treatment. Diagnostic tests were available for those with suspected TB. Those who tested positive started treatment immediately. **Table 4** summarizes barriers and facilitators to 3HP implementation at the clinic.

> “We began with Isoniazid in 2017 when the government began giving us doses. So, from 2018 to 2019, we ensured that all clients are put on isoniazid prophylaxis. The challenge was the shortage of doses because the government would give us about 1,000 doses, and yet we have 6,000 clients so we prioritized who needs it more than the other.” (Female HCP, interview 03).

**Table 4.**
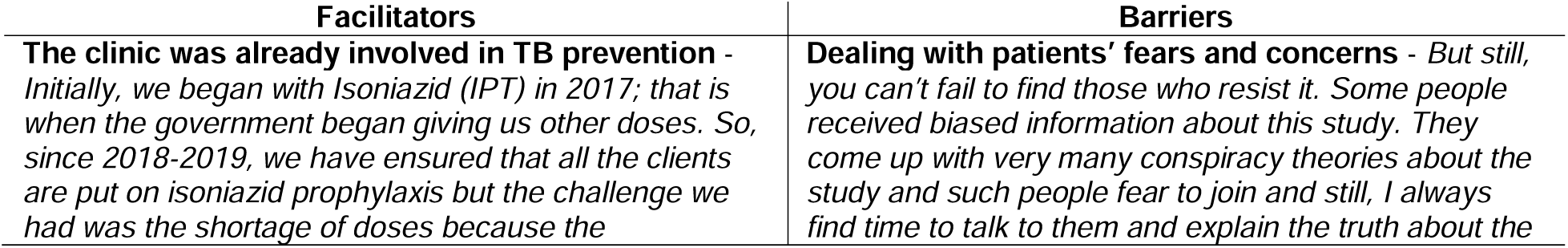
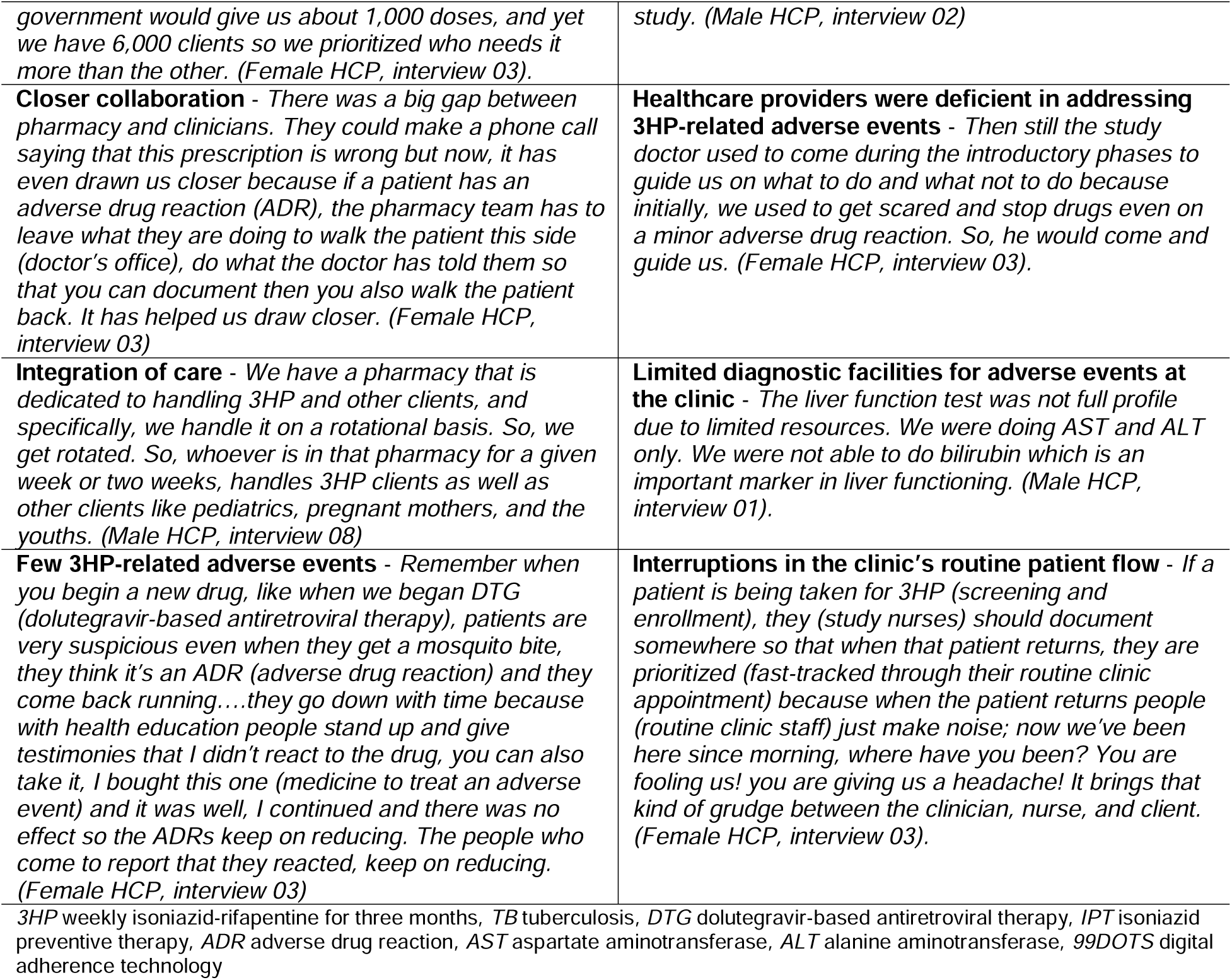
Facilitators and barriers to 3HP implementation at the Mulago ISS Clinic.

Integrating 3HP delivery into the clinic’s daily operations ensured accurate treatment. The clinic streamlined pharmacy-only visits for stable PLHIV, who receive ART refills at a dedicated pharmacy window. The follow-up visits for the treatment of the study participants were conducted similarly. To reduce the workload at the pharmacy, an extra pharmacy technician was hired and included in the team of pharmacy technicians. However, they were not exclusively assigned to 3HP study operations. All pharmacy staff were trained to provide services related to 3HP, and they were rotated every two weeks to ensure their skills were up-to-date. As a result, HCPs did not experience an increased workload due to the introduction of 3HP as they initially anticipated.

> “I would get a maximum of three patients a day reporting 3HP-related side effects, and that is on a few days, but most of the days, I got one patient for 3HP. So, it wasn’t common that patients were presenting with side effects.” (Male HCP, interview 01).

> “We have a pharmacy dedicated to handling 3HP and other clients; specifically, we handle it rotationally. So, we get rotated. So, whoever is in that pharmacy for a given week or two weeks handles 3HP clients and other clients like pediatrics and pregnant mothers, and the youths.” (Male HCP, interview 08).

Concerns among HCPs regarding potential 3HP drug-related side effects were initially reported. However, the small number of serious adverse events helped to alleviate their fears.

> “Remember when you begin a new drug, like when we began DTG (dolutegravir), patients are very suspicious even when they get a mosquito bite, they think it’s an ADR (adverse drug reaction), and they come back running….they go down with time because with health education people stand up and give testimonies that I didn’t react to the drug, you can also take it, I bought this one (medicine to treat an adverse event), and it was well, I continued, and there was no effect so the ADRs keep on reducing. The people who come to report that they reacted (to the new medicine) keep on reducing.” (Female HCP, interview 03).

3HP delivery reportedly promoted closer collaboration between clinicians (doctors and clinical officers) and pharmacy technicians. This teamwork enhanced fidelity to 3HP implementation:

> “…There was a big gap between pharmacy and clinicians. They could make a phone call saying that this prescription is wrong, but now, it has drawn us closer because if a patient has an adverse drug reaction, the pharmacy team has to leave what they are doing to walk the patient this side (doctor’s office), do what the doctor has told them so that you can document and then walk the patient back. It has helped us draw closer.” (Female HCP, interview 03).

#### Barriers to implementation

PLHIV initially hesitated to accept 3HP due to safety concerns, but they consulted their trusted HCPs who eventually convinced them to try it.

> “You know, people are always worried about new things; initially, they have this anticipation that people want us dead. So, each time you bring something new, they are like, are they going to kill us? But you have to convince them that we are not going to kill them, all will be well, and then even some clinic staff had to take these drugs (3HP) to build more confidence in the clients. It helped us build more confidence and take the drugs better.” (Female HCP, interview 03).

During the initial stage of participant recruitment, HCPs felt that they were not adequately equipped to manage adverse events associated with 3HP treatment for PLHIV. Therefore, they tended to be cautious and would discontinue 3HP treatment for PLHIV who experienced adverse events. However, as their confidence grew, clinicians became more willing to give PLHIV a chance to recover from adverse events and continue their treatment. The COVID-19 pandemic further complicated adverse event management for PLHIV on 3HP treatment.

> “There was a season when we got a lot of suspected pulmonary embolism, it scared us a lot, and patients were told to do the cardiac ECHO and electrocardiogram (ECG) tests by themselves; they had to do the CT scan (Chest) by themselves, and they could not afford it. We lost interest somewhere, but when the cases went down, we picked up with time. I think it was due to COVID-19 because COVID-19 was causing those emboli, and since these people were on 3HP, we worried that maybe it is that 3HP that was causing the emboli.” (Female HCP, interview 03).

During the enrolment process, patients were required to undergo certain tests that were not part of the clinic’s routine procedures. This led to disruptions in the patient flow and caused delays. In addition, some patients experienced side effects that required further examination outside of the clinic. Unfortunately, this was often unaffordable for many patients. As a result, it made decision-making regarding the discontinuation of 3HP treatment more complicated. In some cases, decisions had to be made without proper diagnoses.

> “The liver function test was not full profile due to limited resources. We were doing AST and ALT only. We were not able to do bilirubin, which is an important marker in liver functioning.” (Male HCP, interview 01).

Some clinicians were uncomfortable prioritizing study participants over other patients, especially when patients were waiting in the queue.

> “When a 3HP patient presented with a complication, there was a way that the doctor was supposed to dig deep to know more about the complication. This would make the doctor spend more time on the 3HP patient while holding up the queue for others because of the fast-tracking of 3HP patients. So other patients would not feel comfortable.” (Male HCP, interview 01).

### Maintenance

After the completion of the study, the national HIV program started providing 3HP treatment. Our research was presented to the national 3HP planning committee, which helped in the nationwide rollout of the treatment. HCPs continued to triage PLHIV for 3HP treatment and offered standardized pre-treatment counseling to eligible individuals. However, there were some notable differences in the routine programmatic delivery of 3HP treatment compared to the trial. PLHIV receiving 3HP programmatically received all 12 doses of medication at once and were instructed to self-administer them weekly from home. They were asked to report any adverse events to clinic staff via telephone calls (not toll-free) or to return to the clinic for assistance if they experienced any adverse events within the 12 weeks. Interventions such as DOT, travel cost reimbursement, and digital adherence monitoring (99DOTS) were discontinued.

## Discussion

In this qualitative evaluation, we found that facilitated delivery strategies were largely effective in overcoming barriers to 3HP acceptance and completion. Pre-treatment counseling enhanced patients’ understanding of the need for TPT, and the less frequent dosing and shorter treatment duration of 3HP enhanced acceptance. Similarly, regardless of delivery method (SAT vs. DOT), weekly dosing reminders, reimbursement of participants’ travel costs for clinic visits, and social support for PLHIV taking 3HP enhanced convenience and engagement, confirming that the facilitation components selected worked as intended and overcame the barriers to treatment completion for most participants. Among the minority who were unable to complete treatment, side effects and COVID-19 pandemic-related travel restrictions were key contributors. Although effective, the facilitation components other than pre-treatment counseling were not sustained following completion of the trial, largely because they (e.g. travel cost reimbursement, digital adherence monitoring with 99DOTS) require extra costs and logistics. Whether similar levels of treatment completion can be maintained without active facilitation requires further investigation.

Our findings confirm that features of the TPT regimen (safety profile, pill burden, treatment duration, and dosing frequency) are critical to acceptance, as has previously been reported elsewhere (23, 24), and that delivery strategies should be tailored to increase convenience, provide flexibility based on changing circumstances, and ensure PLHIV feel connected to health workers. When appropriate facilitation components addressed these issues, both DOT and SAT were highly acceptable delivery strategies. Participants taking 3HP by DOT emphasized the benefits of having treatment closely monitored by a health worker and the lower risk of inadvertent HIV status disclosure with facility- vs. home-based treatment. Participants taking 3HP by SAT found it helpful to have the ability to self-determine when and where to take treatment. This was particularly helpful for those who were employed or lived far from the clinic. In the patient choice strategy, some PLHIV found the ability to choose and switch between DOT and SAT critical based on their changing circumstances. Our study suggests that both DOT and SAT are viable options for 3HP delivery. The choice between the two methods depends on factors such as proximity to the health facility, ability to self-administer medication once a week, and living situation. Cost and convenience should also be considered when deciding which delivery method to use.

We found that high intervention adoption and implementation fidelity were made possible by training, close collaboration among HCPs, and the integration of the intervention into routine care. This reduced the disruption of routine practices in the clinic. Education prior to participant enrollment alleviated the initial concerns about 3HP safety and empowered HCP to address 3HP-related adverse events. Similarly, Muddu et al. (2023) observed that HCP training and intervention integration into routine care enabled the adoption and implementation of an integrated HIV-hypertension intervention in the same setting (25); whereas Chisare et. al (2021) reported that HCP buy-in and strong collaborative capacity enabled 3HP adoption and implementation in four health care facilities in Zimbabwe in 2020 (26).

Our study had some limitations. First, the study was conducted in a single urban HIV clinic and involved participants who had high research literacy and HCPs who were already familiar with TPT implementation using IPT. This may have made it easier for participants to accept, adopt, and implement 3HP and the results may only apply to similar HIV clinics. Second, we did not interview the four participants who did not initiate 3HP upon enrollment. As a result, we may have missed out on their unique perspectives on why they did not accept 3HP.

## Conclusions

Using an established implementation science framework, we demonstrated that facilitated 3HP delivery is highly acceptable, effective, and feasible. To overcome initial concerns about the safety of 3HP among PLHIV and HCPs, counseling and training are essential. Additionally, active facilitation to minimize costs, increase convenience, and enhance connectedness to HCPs are critical for successful implementation. However, the expenses associated with 3HP facilitation may pose a challenge to wider scale-up, and further studies are required to assess the costs and efficiency of de-escalated facilitation strategies.

## Supporting information

**S1 Checklist. Consolidated Criteria for Reporting Qualitative Research (COREQ)**

## Data Availability

All data produced in the present study are available upon reasonable request to the authors with the necessary editing to preserve participant anonymity.

## Acknowledgments

The authors are grateful to all the administration, staff, and clients at the Makerere University Joint AIDS Program (MJAP), Mulago ISS clinic for their time and participation in the study. We also thank the Infectious Diseases Research Collaboration (IDRC) for supporting the study.

## Authors’ contributions

**Conceptualization:** DP, CAB, PN, RB, MRK, MAH, PPJP, NK, AK, DWD, AC, FCS, ARK

**Data curation:** AM, JRK, JLK, FW, AN, LA, JN, LTK, BO, ARK

**Formal analysis:** AM, JRK, FCS, ARK

**Funding acquisition:** DWD, AC, FCS

**Investigation:** JRK

**Methodology:** AM, JRK, JLK, PN, MRK, MAH, PPJP, NK, AK, DWD, AC, FCS, ARK

**Project administration:** AM, JLK, PPJP, NK, AK, DWD, AC, FCS, ARK

**Supervision:** AM, JLK, DWD, AC, FCS, ARK

**Validation:** FCS, ARK

**Visualization:** AM, JRK, JLK, YB, AS, HEA, RB, MRK, MAH, DWD, AC, FCS, ARK

**Writing – original draft:** AM, JRK, JLK, DWD, AC, FCS, ARK

**Writing – review & editing:** AM, JRK, JLK, FW, AN, LA, JN, LTK, BO, YB, DP, AS, CAB, HEA, PN, RB, MRK, MAH, PPJP, NK, AK, DWD, AC, FCS, ARK

## Notes

### Competing Interest Statement

The authors have declared no competing interest.

### Clinical Trial

ClinicalTrials.gov (NCT03934931) Registered 2nd May 2019 https://clinicaltrials.gov/study/NCT03934931?id=NCT03934931&rank=1

### Clinical Protocols

https://link.springer.com/article/10.1186/s13012-020-01025-8

### Funding Statement

This study was supported by a grant from the US National Heart, Lung and Blood Institute: NIH/NHLBI R01HL144406 (AC and DD). The funder had no role in the study design, data collection and analysis in the preparation of the manuscript or decision to publish.

### Author Declarations

This study was approved by the School of Public Health Research Ethics Committee at the Makerere University College of Health Sciences (Kampala, Uganda), the Uganda National Council for Science and Technology (Kampala, Uganda), and the University of California San Francisco Institutional Review Board (San Francisco, CA, USA).

